# Is a fair comparison between doping tested and doping untested powerlifters possible?

**DOI:** 10.1101/2021.05.07.21256806

**Authors:** Anna Khudayarov

## Abstract

**Purpose:** Powerlifting is clearly divided into doping tested amateur competitions and doping untested pro competitions with pro competition money prizes ranging from a few thousand USD to over $120 000 USD in a single competition. However, the results of both pro and amateur powerlifting competitions are then collected and put into the same result database (although there is the possibility to filter the results using doping control status). The powerlifting results are compared by scores to have a fair comparison between different weight class athletes. This study plays with the thought of comparing doping tested and doping untested athletes, and creates a coefficient for the comparison.

**Methods:** The powerlifting results (noted in kg) of the 10 top ranked athletes per weight class and separately for men and women, and for doping tested and doping untested categories were collected from the openpowerlifting.org database. A weight adjusted model was fit to these results separately for men and women with the doping control status used a binary factor.

**Results:** Doping control status was a significant factor when modelling the powerlifting results, p<0.001 for both men and women. Men’s doping untested results were 1.0725 times as large as the doping tested results. Women’s doping untested results were 1.1208 times as large as the doping tested results. These separate and precise factors could be used as coefficients for scoring doping tested and doping untested athletes’ powerlifting results. Moreover, the coefficients approximate the effect of doping on an elite level athlete’s results.

## INTRODUCTION

Powerlifting is a sport that combines the bench press, squat and deadlift. The powerlifting result is the total of these events. Powerlifting is divided into equipped (using support suits) and RAW (no suits). RAW powerlifting is the most popular form of powerlifting with nearly 70 000 athletes Worldwide in 2018 (www.openpowerlifting.org). RAW powerlifting (with knee wraps) is also a form of powerlifting that has money prize competitions. The largest annual Pro Raw powerlifting competition is the US Open, with up to $120 000 USD in prize money.

There are more than 50 separate powerlifting federations, with minimal differences in their rules. The main amateur federations have doping tests. Pro competitions are not necessarily connected to any federation, and do not have doping control. All official competition results are collected for the same database and ranking list at openpowerlifting.org. These results can be filtered by the different forms of powerlifting, doping control status, and weight class.

A powerlifting result is highly dependent on an athlete’s body weight. Several scoring systems have been used to compare different weight class athletes, and choose the winners of the competitions. The classical theoretical models represent allometric modelling (modelling of the relationship of body size to physiological phenomena).[1] The allometric dependence of weight lifting results and body weight was first shown 1956 by Lietzke.[2] The ideal allometric model for strength is y=a*x^2/3^, as strength is thought to be directly proportional to the cross-sectional area of muscle (square of length), and mass is the cubic of length, so strength is proportional to mass power 2/3. The currently used scoring systems are more statistically than theoretically derived.[3–7] There are separate coefficients for masters and teens’ classes, and these can be used together with any of the other scoring systems.

Anabolic steroids were tested for the first time at the Olympic Games 1976, and they have ever since constituted the vast majority of positive doping cases.[8] Anabolic androgenic steroids are the main drugs of abuse in strength sports, especially in untested pro powerlifting. It is not known, and would be unethical to test, how much these substances exactly benefit elite level athletes.[9]

As powerlifting already uses scoring systems, it would not be a large new step to include the coefficients so as to compare doping tested and doping untested athletes to each other - for ranking list, not for competition use. This current study calculates these coefficients, separately for men and for women.

## MATHERIALS & METHODS

For this study, 10 top-ranked athletes’ RAW (with or without knee wraps) powerlifting results per weight class for every traditional weight class (−52 kg, -56 kg, -60 kg, -67.5 kg, -75 kg, - 82.5 kg, -90 kg, -100 kg, -125 kg, -140 kg, and +140 kg for men, and -44 kg, -48 kg, -52 kg, - 56 kg, -60 kg, -67.5 kg, -75 kg, -82.5 kg, -90 kg, and +90 kg for women) were collected from the openpowerlifting.org current All-Time ranking, i.e., separately for men and women, and also separately for doping tested (using the filter “All doping tested athletes”) and all athletes (no filter, thus here called “untested”). The doping tested results were collected on 3 March 2019, and the untested results were collected on 27^th^ February 2019, from the ranking list. That list is updated on a daily basis.

By the Finnish Law of Medical Research (488/1999), a study using public data does not need approval by the local ethics committee. This study meets international ethical standards.

An allometric model was fit separately to the doping tested and untested results and to the men and women’s results. Then, as the 95 % confidence intervals of the exponents overlapped, an allometric model using doping test status as a binary factor was fitted to the results, separately for men and women (the men’s and women’s exponent for 95 % confidence intervals did not overlap). An allometric model was chosen, because in that model, a factor produces a coefficient rather than an intercept, as in polynomial regression.

The analyses were done using the freely downloadable Jamovi software on a Windows PC. The plots were created using Microsoft Excel.

## RESULTS

The doping tested and doping untested RAW powerlifting results had clearly different slopes, the difference between the groups growing toward the larger weight classes, as shown in **Figure 1A-B**. An allometric model was fit first separately to the doping tested and doping untested results, and also to the men’s and women’s results, using *ln* transformed variables. Thus, the formulas were:

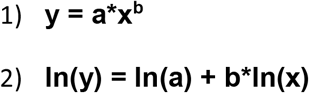

**Figure 1A-B.**
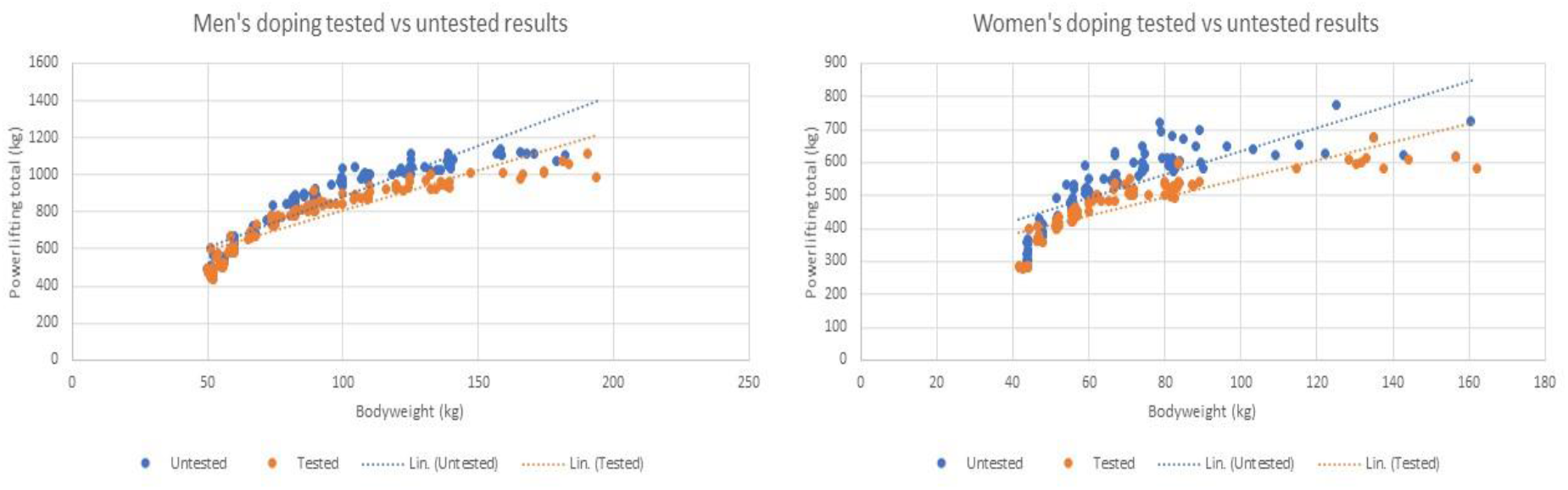
Men and women’s doping tested and doping untested RAW powerlifting results shown in dot plots. The doping tested and the doping untested results cannot be described using the same slope.

The estimates and their 95 % confidence intervals are collected in **Table 1**. The 95 % confidence intervals of the exponent (b) of the doping tested and the doping untested populations overlapped in both the women and men’s data, so a common model could be fit for the doping tested and doping untested results. Women’s doping tested and men’s doping untested groups’ 95 % confidence intervals of the exponent (b) did not overlap, so the model was fit to men and women separately.

**Table 1.** The estimates and their 95 % confidence intervals and the p values for the fit allometric models separately for men and women, and also for doping tested and doping untested athletes.

| $\ln(y) = \ln(a) + b \cdot \ln(x)$ | Estimate | 95 % Confidence interval | p value |
| --- | --- | --- | --- |
| Men, doping tested |  |  |  |
| *a* | 3.915 | 3.719 - 4.112 | <0.001 |
| *b* | 0.608 | 0.564 - 0.652 | <0.001 |
| Men, doping untested |  |  |  |
| *a* | 3.590 | 3.389 - 3.791 | <0.001 |
| *b* | 0.697 | 0.652 - 0.742 | <0.001 |
| Women, doping tested |  |  |  |
| *a* | 3.808 | 3.526 - 4.089 | <0.001 |
| *b* | 0.553 | 0.486 - 0.621 | <0.001 |
| Women, doping untested |  |  |  |
| *a* | 3.644 | 3.330 - 3.959 | <0.001 |
| *b* | 0.620 | 0.545 - 0.695 | <0.001 |

An allometric model using the doping test status (D) as a binary factor (1 for the doping tested, 0 for doping untested) was fit to the data for men and women separately. These formulas were:

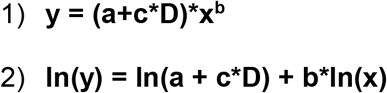

The estimates and their 95 % confidence intervals are shown in **Table 2**. The fitted functions as plotted against the data are shown in **Figure 2A-D**. The residual plots are shown in **Figure 3A-B**. The doping test status was a significant (p<0.001) factor in both the men and women’s models. The coefficients of correlation were high at R^2^=0.876 for men, and R^2^= 0.749 for women.

**Table 2.** The estimates and their 95 % confidence intervals and p values for the fit allometric models shown separately for men and women, doping test status (D) included as a binomial factor.

| $\ln(y) = \ln(a + c \cdot D) + b \cdot \ln(x)$ | Estimate | 95 % Confidence interval | | p value |
| --- | --- | --- | --- | --- |
|  |  | Lower | Upper |  |
| Men |  |  |  |  |
| a | 3.7591 | 3.6174 | 3.9007 | <0.001 |
| b | 0.6509 | 0.6192 | 0.6826 | <0.001 |
| c | -0.0700 | -0.0927 | -0.0472 | <0.001 |
| Women |  |  |  |  |
| a | 3.735 | 3.525 | 3.9443 | <0.001 |
| b | 0.585 | 0.535 | 0.6345 | <0.001 |
| c | -0.114 | -0.146 | -0.0822 | <0.001 |

**Figure 2A-D.**
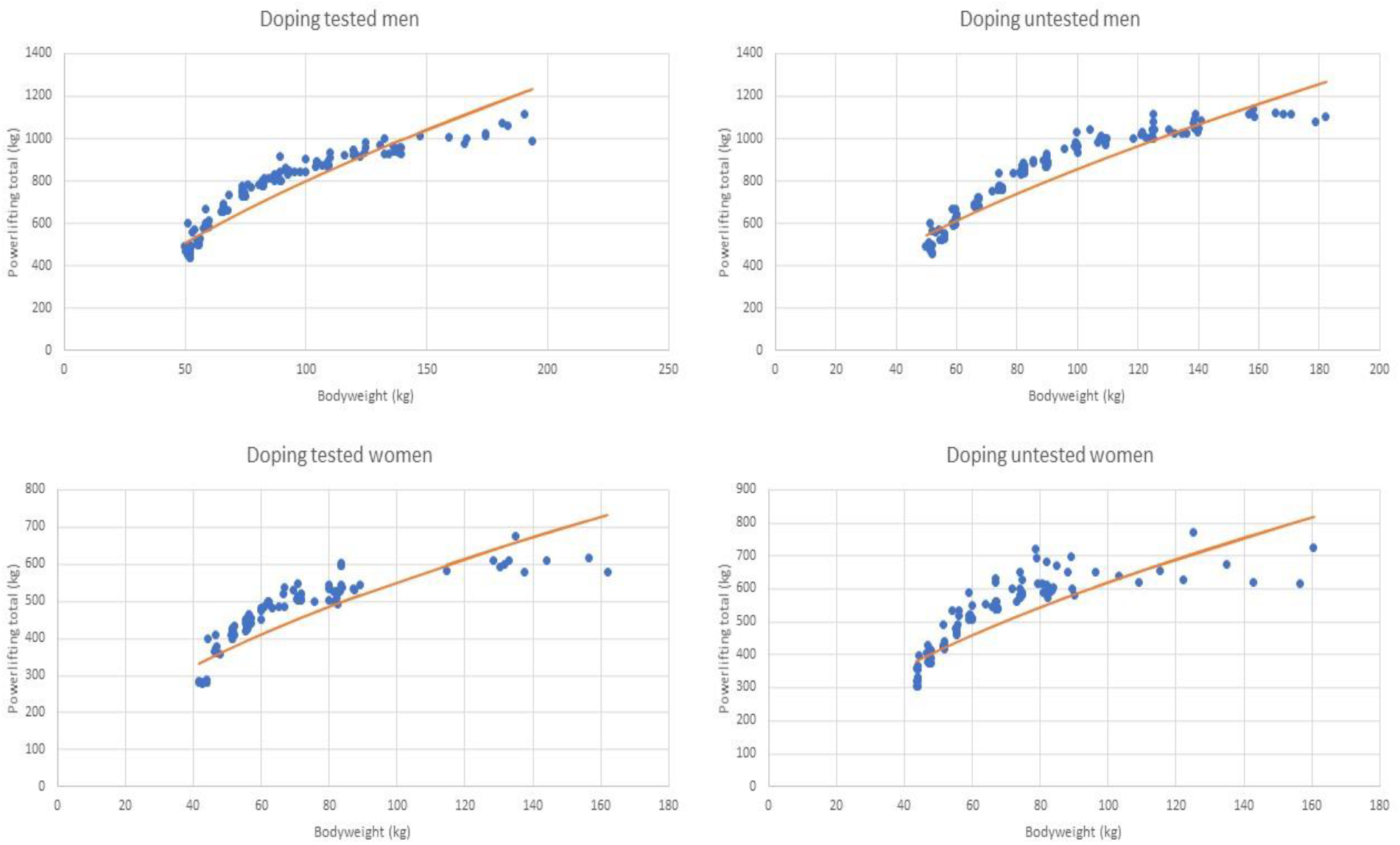
The fit allometric prediction functions plotted against the powerlifting results.

**Figure 3A-B.**
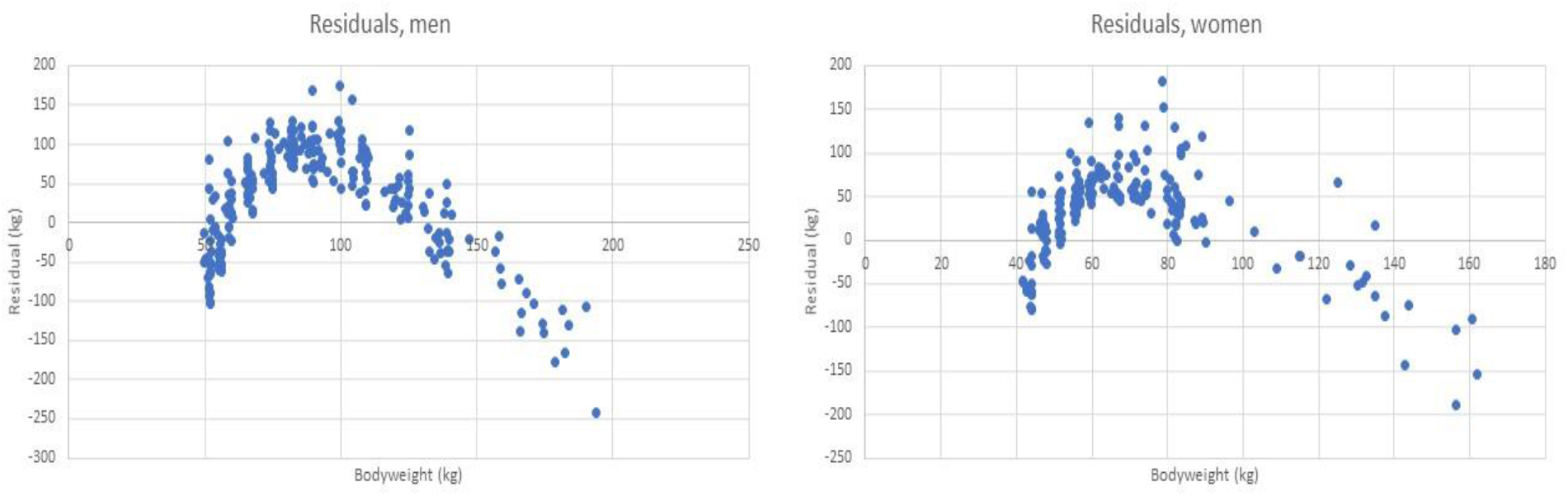
Residuals of the models fit above, using doping test status as a binomial factor. The residuals are not normally distributed, but positive for the middleweight classes, and negative for the extremes of body weight. An allometric model is not a suitable system for adjusting powerlifting results by body weight when scoring competition results, but it is an acceptable tool for calculating estimates of binomial factors.

These final formulas were:

Doping tested men:

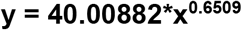

Doping untested men:

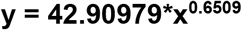

Doping tested women:

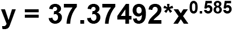

Doping untested women:

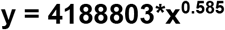

According to the weight-adjusted model created above, the men’s untested results were 1.0725 times as large as the men’s doping tested results; the women’s untested results were 1.1208 times as large as the women’s doping tested results. These ratios could be used as coefficients for scoring powerlifting results when comparing doping tested and untested athletes to each other.

## DISCUSSION

The coefficients calculated here do not represent the effect of doping directly, but rather the effect of doping tests, as there is no sure information about who of the athletes really use doping. There is clearly also doping use among the doping tested athletes, as there are also positive doping tests. Also, the “untested” group here represented non-sorted ranking list results, so there were also some doping tested athletes in this group, especially in the lightweight classes.

There are also other factors that contribute to the difference between the doping tested and the untested powerlifting results, as the different powerlifting federations do have slightly different rules. The pro powerlifting competitions mostly hold their weigh-ins 24 hours before competition, whereas amateur competitions mostly have 2-hour weigh-ins before the actual competition. There are pro competitions also with 2-hour weigh-ins, and amateur competitions with 24-hour weigh-ins. Amateur competitions mostly do not allow the use of knee wraps in RAW powerlifting category, whereas pro money competitions do present RAW powerlifting with knee wraps. There are also exceptions to this rule. In this current study, RAW was considered with and without knee wraps, and thus both results were included in this study population, where they belonged to the top 10 results of each weight class. There are more high-weight class athletes in doping untested powerlifting, and more lightweight class athletes in doping tested powerlifting. The allometric model used in this study corrects the results for weight. Overall, different factors were contributing to the difference between the doping tested and doping untested athlete powerlifting results evening each other out, so the coefficients calculated here also approximate the effect of doping on an athlete’s results.

Allometric models are the theoretical choice of models for adjusting powerlifting results by body weight. However, they have been shown to favour middleweight class athletes in a mathematically biased way, as the residuals of the fitted prediction curve tend to be positive for the middleweights, and negative at both the extremes of weight, as was the case also with the models fitted in this study and shown in **Figure 3A-B**. So allometric modelling is not the choice of model for producing new scoring systems to adjust powerlifting results by body weight. Here allometric modelling was chosen as the simplest model for calculating coefficients using binary (0-1) factors in the model.

The openpowerlifting.org ranking list of powerlifting results uses a polynomial adjustment function for scoring as a basic setting. A polynomial model could not be used in this study, as the difference between the powerlifting results for the tested and untested athletes was clearly proportional to the powerlifting result – or the body weight, with larger difference towards the heavyweight classes. It might also be that athletes using a large amount of anabolic doping substances simply grow larger in body weight, which results in heavier doping use in the heavyweight classes. Further, the anabolic substances might help athletes surpass the physiological limit of a maximal lean body mass. Thus, the fat-free body weight of the super heavy athletes might be considerably higher in the doping untested group compared to the doping tested group, whereas there is no such difference in the lightweight classes.

The exponents (b) calculated in this study are of the same order of magnitude as the earlier findings (1, 2, 5). The women’s exponent is lower than the men’s, most likely because of a larger body fat percentage of high weight class women, as strength correlates to lean body mass rather than to full body mass. The weight adjustment functions that were fitted in this study are not meant for use in the weight adjustment of competition results. Their only purpose was to calculate the coefficients for comparing the doping tested to the doping untested groups.

Interestingly enough, having the possibility to choose between doping tested and doping untested competitions or federations might make powerlifting a fair field for sport. Those athletes who are willing to use doping substances are more likely to choose untested competitions, rather than secretly use doping substances in competitions, where they are prohibited and thus tested. The discrepancy between doping tested and doping untested athletes is mostly associated with result ranking lists, as there is no need to compare tested and untested athletes in a single competition. Currently the overall powerlifting ranking list is listed by weight adjusted scores. It could be possible, however, to add extra coefficients, so as to compare doping tested and doping untested powerlifting results in the score ranking setting. Even if not used in any official setting, the possibility to compare doping tested and doping untested athlete’s results might both promote friendly rivalry and motivate all athletes to achieve better results.

The coefficients calculated here, namely 1.0725 for men and 1.1208 for women, might be a useful tool to use for comparing the powerlifting scores of doping tested and doping untested athletes. In this case, being doping tested would give extra points. These coefficients also approximate the effect of doping on an athlete’s results, although the coefficient cannot be used to predict the effect of doping on the individual level. Here, the estimated effect of doping on a male athlete’s powerlifting result was 7.25 %, and on a female athlete’s result 12.08 %.

## Data Availability

The data was collected from openpowerlifting.org database February-March 2019. The full can be provided upon request.

https://www.openpowerlifting.org/

## ACKNOWLEDGEMENTS

We thank openpowerlifting.org administration for keeping this open access powerlifting result database up to date.

## STATEMENTS

The corresponding author is a competitive powerlifter, and any scoring or ranking system changes will affect her sports career. There are no other conflicts of interest to disclose.

